# Exploring the Efficacy and Potential of Large Language Models for Depression: A Systematic Review

**DOI:** 10.1101/2024.05.07.24306897

**Authors:** Mahmud Omar, Inbar Levkovich

## Abstract

**Background and Objective:** Depression is a substantial public health issue, with global ramifications. While initial literature reviews explored the intersection between artificial intelligence (AI) and mental health, they have not yet critically assessed the specific contributions of Large Language Models (LLMs) in this domain. The objective of this systematic review was to examine the usefulness of LLMs in diagnosing and managing depression, as well as to investigate their incorporation into clinical practice.

**Methods:** This review was based on a thorough search of the PubMed, Embase, Web of Science, and Scopus databases for the period January 2018 through March 2024. The search used PROSPERO and adhered to PRISMA guidelines. Original research articles, preprints, and conference papers were included, while non-English and non-research publications were excluded. Data extraction was standardized, and the risk of bias was evaluated using the ROBINS-I, QUADAS-2, and PROBAST tools.

**Results:** Our review included 34 studies that focused on the application of LLMs in detecting and classifying depression through clinical data and social media texts. LLMs such as RoBERTa and BERT demonstrated high effectiveness, particularly in early detection and symptom classification. Nevertheless, the integration of LLMs into clinical practice is in its nascent stage, with ongoing concerns about data privacy and ethical implications.

**Conclusion:** LLMs exhibit significant potential for transforming strategies for diagnosing and treating depression. Nonetheless, full integration of LLMs into clinical practice requires rigorous testing, ethical considerations, and enhanced privacy measures to ensure their safe and effective use.

**A visual abstract:** 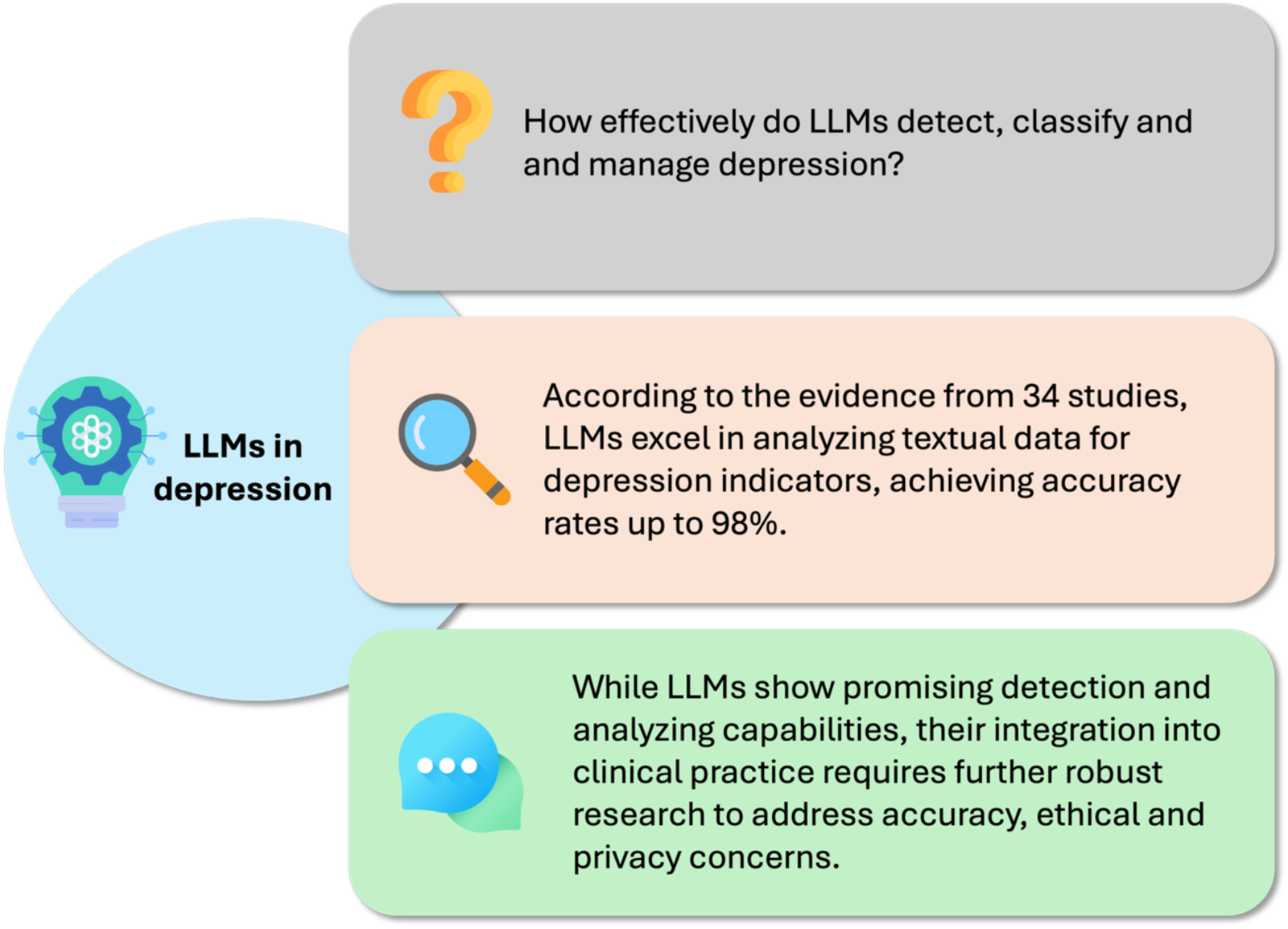

## Introduction

Mental health has emerged as a significant concern in modern society. Specifically depression is a major challenge for global healthcare due to its prevalence and impact on quality of life (1,2). Ongoing advancements in artificial intelligence, particularly large language models (LLMs), have revolutionized the diagnosis and treatment of depression (3–5). Advanced versions of these models, such as ChatGPT and Claude, leverage their extensive linguistic capabilities to facilitate early diagnosis and intervention, which are crucial for improving mental well-being (5–7).

Traditional treatment systems often face obstacles such as high costs and limited resources, which delay the provision immediate support for mental health issues (8,9). The inherent potential of LLMs offers a promising solution to these obstacles by enhancing accessibility and overcoming geographical, financial, and stigma barriers, thereby facilitating personalized treatment and management of depression (10–12).

According to healthcare professionals, LLMs have demonstrated effectiveness in preliminary assessments and treatment times (3,13), yet they cannot replace human therapists. Instead, they can serve as a supplementary tool that integrates human clinical insights to improve therapeutic processes (14,15). Nevertheless, several challenges to the efficacy of using LLMs in psychiatry still remain, including bias and the nascent stage of research (14,15).

This review aimed to provide a comprehensive analysis of the role of LLMs in enhancing the understanding and treatment of depression, thus addressing a gap in systematic reviews focusing on the impact of AI in this area.

## Methods

### Registration and Protocol

This systematic literature review was registered with the International Prospective Register of Systematic Reviews (PROSPERO) under the registration code CRD42024539720 (16). Our methodology adhered to the Preferred Reporting Items for Systematic Reviews and Meta-Analyses (PRISMA) guidelines (17).

### Search Strategy

Between January 2018 and March 2024 we conducted a comprehensive search of key databases, including PubMed, Embase, Web of Science, and Scopus. To enhance our search, we also used reference screening to identify additional relevant studies. Precise Boolean search strings were meticulously crafted for each database, with a focus on the integration and impact of LLMs on depression analytics. Details of the specific Boolean strings used are provided in the Supplementary Materials.

### Study screening and selection

Given the rapidly evolving nature of LLM research, our review encompasses original research articles, preprints, and full conference papers (18). Review papers, case reports, commentaries, protocol studies, editorials, and publications not written in English were excluded, enabling us to encompass a broader spectrum of the latest advancements in the field. For the initial screening, we used the Rayyan web application (19). Initial screening and study selection were conducted according to predefined criteria and independently carried out by two reviewers (MO and IL). Discrepancies were resolved through discussion.

### Data Extraction

The researchers MO and IL conducted data extraction using a standardized format to ensure consistent and accurate data capture. The format included details such as author, publication year, type of study, sample size, data type, task type, specific task, model used, results, numeric metrics, conclusions, and limitations. Any discrepancies in data extraction were resolved through discussion and a third reviewer was consulted when necessary.

### Risk of Bias Assessment

To ensure a thorough evaluation of the included studies, we used three distinct tools, each tailored to a specific study design within our review. The ROBINS-I tool was employed for interventional studies assessing LLMs in applications such as management, prescription guidance, and clinical inquiry responses (20). The QUADAS-2 tool was used for diagnostic studies that compared LLMs with physicians or a reference standard for diagnosing and detecting depression (21). Finally, the PROBAST tool was utilized for the remaining studies, which involved the use of LLMs to predict and classify the presence and type of depression from extensive datasets, without direct comparison to reference standards (22). This multitool approach allowed us to address the diverse methodologies and applications considered in the reviewed studies, ensuring a comprehensive and tailored risk-of-bias assessment.

## Results

### Search Results and Study Selection

Our systematic search targeted studies published since 2018, the year the first public LLM was initiated (23). We began by excluding non-relevant publication types, such as reviews, letters, editorials, and comments. The initial search across four databases yielded 449 articles: PubMed (99), Embase (127), Scopus (76), and Web of Science (147). After 76 duplicates were removed, a total of 373 articles remained. Further screening of the titles and abstracts led to the exclusion of another 298 articles, yielding 75 studies for full-text evaluation. Of these, we excluded 19 that did not utilize LLMs and 31 that did not directly evaluate their impact, resulting in 25 studies that met all inclusion criteria. An additional nine studies were included through reference checking and snowballing techniques. **Figure 1** shows the PRISMA flowchart depicting the visual representation of the screening process.

**Figure 1:**
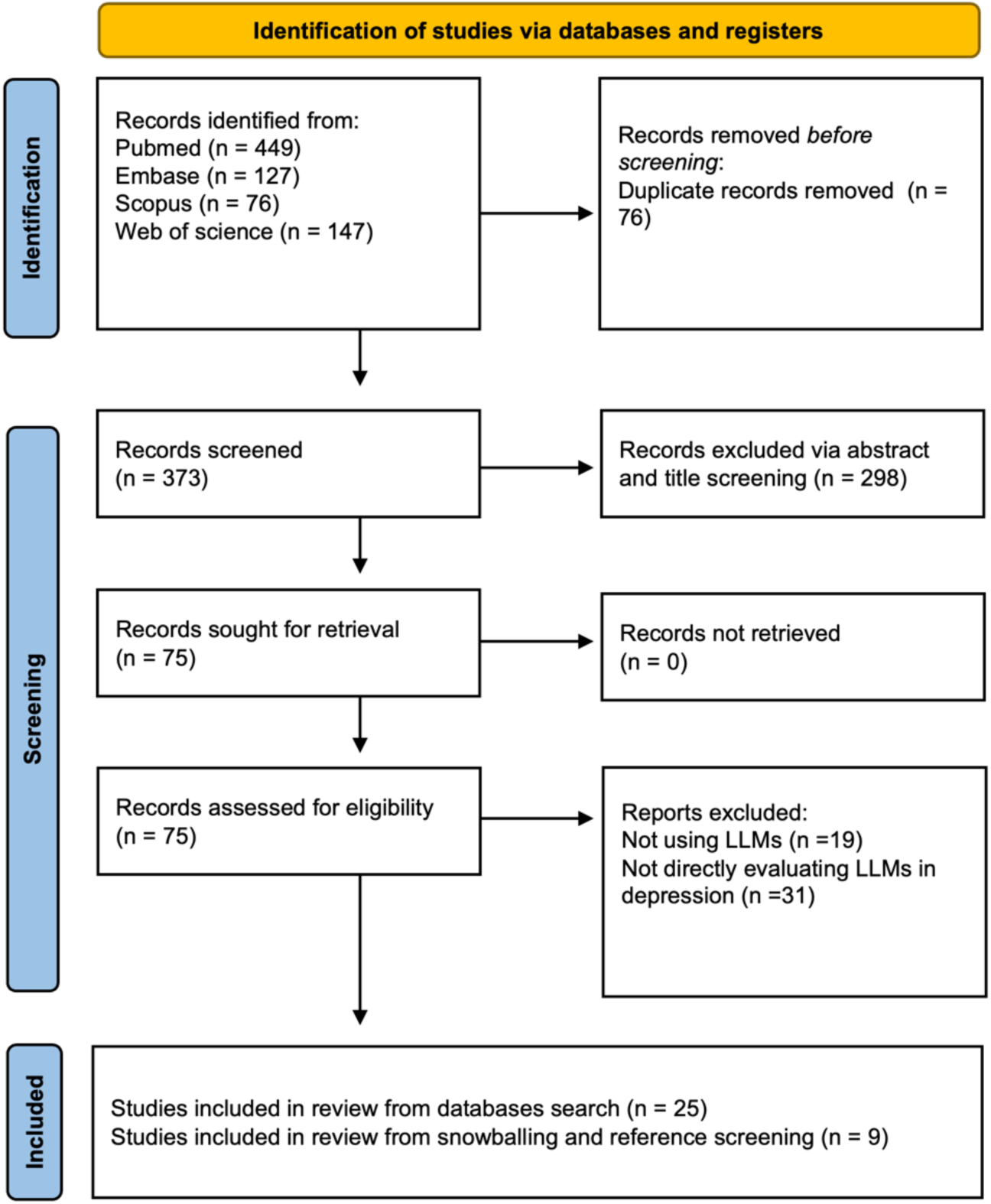

### Overview of Included Studies

As noted, the systematic review included 34 studies published between February 2019 and March 2024 that investigate the application of LLMs in various aspects of depression research (3,7,24,24–54). These studies encompassed a wide variety of sample sizes, ranging from as few as 25 to over 632,000, and utilized data types ranging from clinical interview transcriptions and electronic health records to user-generated content on social media platforms (**Table 1**).

The tasks explored in these studies focused primarily on the detection and classification of mental health conditions. Specifically, LLMs were employed in 13 studies to detect signs of depression or mental health risks using both clinical data and online platforms. Another 15 studies used LLMs to classify depression severity or to differentiate between various mental health conditions using clinical scales and analyses of unstructured electronic health records. Six additional studies assessed the capability of LLMs to recommend treatment strategies or manage depressive episodes, highlighting their potential utility in clinical decision-making (**Figure 2**).

**Figure 2:**
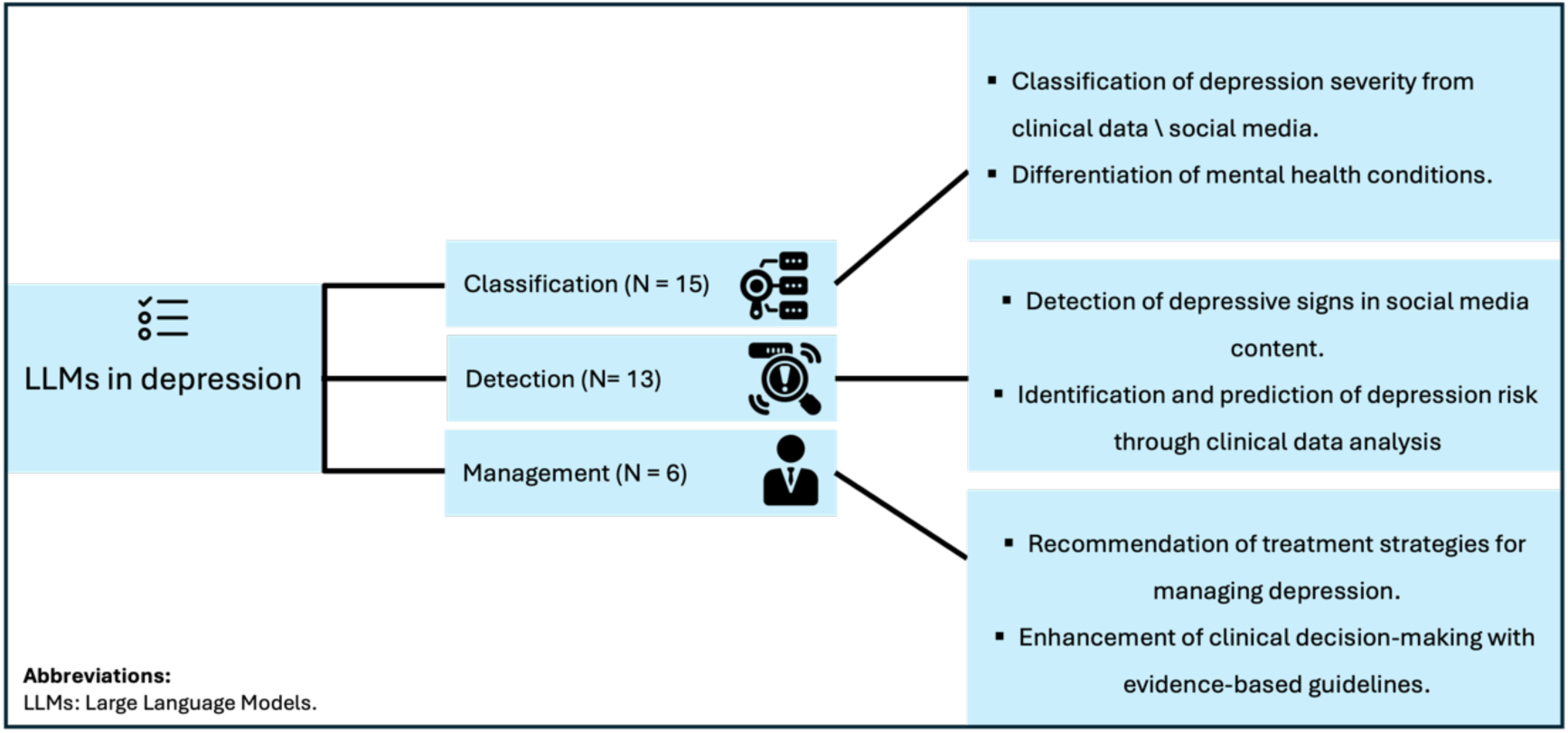

A variety of LLMs were employed in these studies, including prominent models such as BERT and RoBERTa and their derivatives, such as DistilBERT and DeBERTa, as well as different iterations of the GPT models. The most commonly used model among the reviewed studies was RoBERTa, which was effectively applied in various contexts to analyze textual data for signs of depression. In one study, for instance, RoBERTa achieved an accuracy rate of approximately 98% when analyzing Twitter data on depressive signs (31). Other models, such as BERT and its variations as well as GPT models, also showed substantial effectiveness across different datasets, particularly in tasks involving the classification of mental health conditions from unstructured data.

### Risk of Bias

Our assessment of the risk of bias across the included studies reveals a nuanced landscape, with variations in the rigor of methodology that reflect the pioneering nature of LLMs research. By employing ROBINS-I, QUADAS-2, and PROBAST, we carefully mapped potential biases and adapted these robust tools to the specific contours of each study. Note that most of the included studies were published in Q1 journals, indicating a high level of scholarly impact, with robust SCImago Journal Rank (SJR) scores, as illustrated in **Figure 3**.

**Figure 3:**
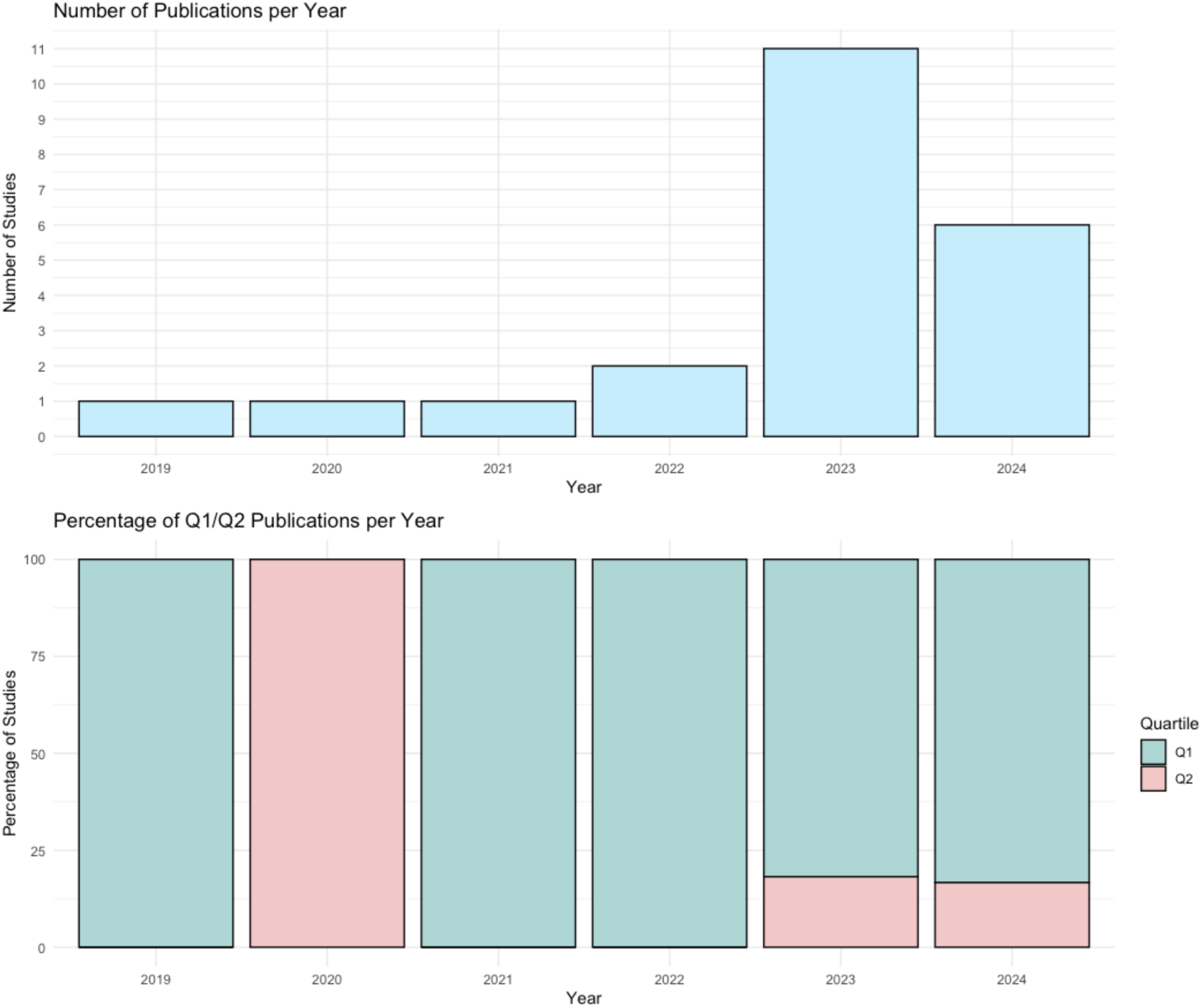

**QUADAS-2 (Table S1):** A synthesis of the QUADAS-2 results revealed that among the studies evaluated, one exhibited a high risk of bias in patient selection—a pivotal aspect influencing the integrity of the findings. Conversely, multiple studies, such as that by Lau et al. (26), successfully navigated these challenges, demonstrating low risk across all QUADAS-2 domains and underscoring their methodological robustness.

**ROBINS-I (Table S2):** Analysis of the ROBINS-I results revealed that the majority of studies achieved a low risk of bias in measurements and outcomes, indicating a trustworthy basis for their conclusions. Nevertheless, nearly one-third of the studies, including those by Levkovich et al. (7), exhibited moderate biases due to confounding factors and participant selection that may have affected the applicability of the results.

**PROBAST (Table S3):** PROBAST assessments indicated a predominance of low-risk in domains related to outcome and analysis across most studies, as exemplified by Hond et al. (50). Still, a notable proportion of the studies encountered high participant-related bias, affecting the generalizability of their conclusions.

### Refining Diagnostic Precision with LLMs

Within the scope of the classification, 15 studies provided a comprehensive picture of the role of LLMs in mental health diagnostics. These tools showed promising results in parsing complex data into depression severity metrics. Lau et al. (26), for example, found that LLMs were superior to traditional methods in predicting depression from interview transcripts. Dai et al. explored the classification potential of BERT-based LLMs across a spectrum of psychiatric conditions, offering a glimpse into the models’ diagnostic acumen (32). Yet as Wan et al. cautioned, the accuracy of LLMs can be hampered by imbalanced datasets and the multidimensional nature of psychiatric symptoms (44).

**LLMs as beacons in detecting depression** were the focus of 13 studies, in which LLMs such as RoBERTa stood out for their ability to sift through social media and clinical data for signs of depression. Bokolo et al. emphasized the adeptness of RoBERTa in mining Twitter for depressive language, showing a high accuracy rate (31). In addition, Owen et al. demonstrated that BERT-based models were able to discern linguistic patterns indicative of depression weeks before a clinical diagnosis (39). Ye this study also points out the intricacies involved in detecting early depression indicators, serving as a reminder that despite their potential, LLMs still must grapple with the nuances of spontaneous human expression and the vast heterogeneity of online discourse (39).

**LLMs in the Landscape of Depression Management.** In the domain of management, six studies illustrate LLMs’ nascent integration into clinical decision-making. Levkovich et al. demonstrate the potential of ChatGPT in generating treatment recommendations, suggesting a possible future role for LLMs to assist in therapeutic strategy planning (7). The work of Sezgin et al. supports the notion that LLMs can provide clinically sound advice, highlighting ChatGPT’s application in providing information on postpartum depression (28). Nevertheless, as noted by Perlis et al., the efficacy and safety of such tools in real-world clinical settings remains to be thoroughly investigated, underscoring the importance of human oversight in the use of LLMs for clinical purposes (29).

The synthesis of results across the reviewed studies revealed a transformative trend, showing that LLM use is ushering in a new era of proactive, precise, and personalized mental health care. Not only do these models enhance the accuracy of depression detection from various text sources, they also advance the capabilities of mental health diagnostics to anticipate and intervene in the early stages of mental health problems. The evidence suggests that integrating LLMs into regular health monitoring systems could significantly improve early detection rates and personalization of treatment strategies, promising a future in which mental health care is more responsive and attuned to individual needs.

Indeed, current literature suggests that LLMs offer versatile avenues for integration into mental health practices, specifically for the detection, management, and classification of depression (**Figure 4**).

**Figure 4:**
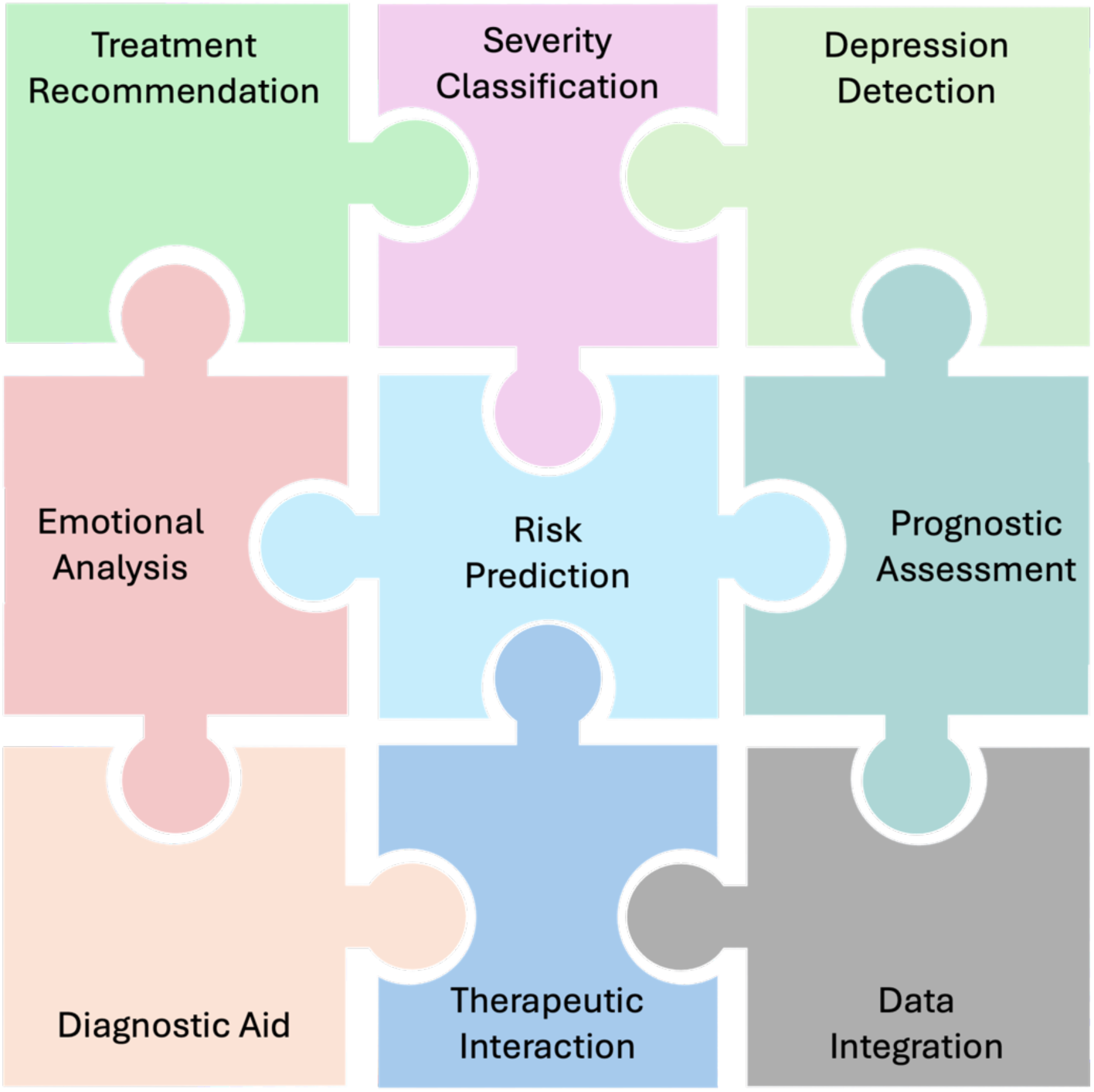

## Discussion

This systematic review examined the efficacy of LLMs in the diagnosis and management of depression, illustrating their potential to revolutionize mental health care. Our analysis identified a critical limitation: the lack of public datasets available for exploring the intersection between depression and artificial intelligence (AI). A survey of 449 datasets over the past six years revealed that only 34 explicitly focused on depression. This scarcity of targeted data significantly hinders comprehensive monitoring and research of depression within the AI domain, underscoring a crucial gap in the resources necessary for advancing our understanding and fostering innovation in this field.

Our findings further demonstrate that LLMs such as RoBERTa are highly effective in rapidly detecting and categorizing signs of depression, achieving accuracy rates as high as 98% in certain instances (31). These models are competent in analyzing texts from both clinical settings and social media platforms, suggesting their potential for facilitating early diagnosis and enabling timely interventions (28). Yet challenges remain, including issues related to data bias and the necessity of human oversight.

These issues emphasize the need for careful integration of LLMs into existing healthcare frameworks to augment rather than replace traditional diagnostic and treatment practices (7,51) (Figure 5).

Our extensive review highlights the rapidly evolving nature of this research field. Most studies prominently feature BERT-based models, underscoring the developmental stage of LLMs in this domain in that newer and potentially more capable models, such as GPT-4 and Google’s Gemini, are less represented. This may suggest that the field is in the nascent stage of adopting cutting-edge LLMs and instead focuses on proven, familiar technologies. Nevertheless, most of the included studies were published in 2023, reflecting growing interest in the most recent developments in the field.

The primary applications of LLMs focused predominantly on the detection and classification of depression from both clinical and social media data. This emphasis underscores the significance of LLMs in enhancing diagnostic processes and in managing and recommending treatment strategies. Our findings resonate with and augment the current literature, as highlighted by Mendez et al. and Omar et al., both of which discuss the vast potential of LLMs in healthcare, particularly in handling large datasets for improving healthcare delivery and patient outcomes (10,55).

In addition to these promising applications, our study also acknowledges critical challenges, such as concerns about data privacy and the need for model transparency, issues that are similarly emphasized in the literature by De Freitas et al. and King et al. (2023) (56,57). These authors critically evaluated the safety and readiness of AI technologies in mental health, pointing to the risks associated with premature deployment and the “black box” nature of AI systems. Their concerns echo our call for cautious integration of these technologies, highlighting the need for robust regulatory frameworks and transparency to mitigate potential risks. These findings also underscore ethical concerns surrounding the widespread use of AI in mental health and highlight significant concerns regarding the impact of AI on human well-being. Among these are the risk of medical errors, potential discrimination that may exacerbate health disparities (58), and the spread of misleading medical information or unverified treatments that could compromise general practitioners’ understanding of medical conditions (59). Despite the potential of AI to enhance medical training through realistic patient scenarios, the risks associated with its misuse remain a critical consideration (60).

Although LLMs showed better results than traditional tools such as machine learning (26,31) and even exhibited capabilities comparable to those of human experts in some cases (3,7), variations in accuracy and output correctness across different tasks persist (7,29). Advanced models such as GPT-4 have been effective in interpreting clinical and unstructured data to manage, detect, and classify depression (26,31,35). However, studies often rely on fictional clinical vignettes, limiting the generalizability of these findings (3,7,29).

While we concur with King et al. that LLMs are not yet ready for full integration into daily psychiatric practice (56), our review suggests that the process of integration has already commenced and is evolving rapidly, with promising outcomes. Our analysis focusing on depression-related studies reveals a more specific and dynamic exploration of LLM applications compared to broader reviews like those by Omar et al., De Freitas et al. and King et al. himself (10,56,57). Given these developments, we advocate continued and expanded research, particularly through more robust methodologies, such as randomized controlled trials and clinical studies. Such an approach can ensure that the integration of LLMs into psychiatric care will be both evidence-based and cautiously implemented, maximizing potential benefits while addressing safety and efficacy concerns.

This systematic review is the most recent and thorough examination of depression-specific applications of LLMS. Furthermore, it employs three distinct tools to provide a comprehensive assessment of risk of bias. Nevertheless, limitations persist both in the included studies and in our review methodology. The primary limitations in the included studies are issues of data imbalance and generalizability of the findings due to the diversity of data types and study settings. Our systematic review was also constrained by the exclusion of non-English studies and the inability to perform a meta-analysis, which was attributed to the heterogeneity of the included studies (61,62). This approach, while necessary to maintain focus and clarity, may overlook valuable insights from broader, multilingual sources and varied research methodologies.

## Conclusion

The field of LLMs in mental health is expanding rapidly, yet it remains somewhat anchored to earlier models such as BERT, indicating a lag in the adoption of the latest technologies, such as GPT-4. Currently, LLMs are invaluable tools for managing unstructured text and monitoring social media, demonstrating their utility in real-time mental health assessments. Nevertheless, they have not yet been fully integrated into daily clinical practice. The application of LLMs in clinical settings and the associated ethical and privacy concerns require further exploration through robust methodologies and clinical trials to ensure their safe and effective use in patient care.

## Supporting information

Tables included in the manuscript

## Data Availability

All data produced in the present work are contained in the manuscript

## Acknowledgment

None

## Supplementary materials

### Specific Boolean strings used for each database

#### PubMed

((“Large Language Model”[All Fields] OR “LLM”[All Fields] OR “Generative Pre-trained Transformer”[All Fields] OR “GPT”[All Fields] OR “GPT-2”[All Fields] OR “GPT-3”[All Fields] OR “GPT-3.5”[All Fields] OR “GPT-4”[All Fields] OR “ChatGPT”[All Fields] OR “Transformer models”[All Fields] OR “BERT”[All Fields] OR “BARD”[All Fields] OR “Gemini”[All Fields]) AND (“Depression”[All Fields] OR “Depressive disorder”[All Fields] OR “Major depressive disorder”[All Fields] OR “Clinical depression”[All Fields] OR “Mood disorder”[All Fields])) AND (2018:2024[pdat]).

#### Embase

(‘large language model’ OR ‘llm’ OR ‘generative pre-trained transformer’ OR ‘gpt’ OR ‘gpt-2’ OR ‘gpt-3’ OR ‘gpt-3.5’ OR ‘gpt-4’ OR ‘chatgpt’ OR ‘transformer models’ OR ‘bert’ OR ‘natural language processing’ OR ‘bard’ OR ‘gemini’) AND (’depression’ OR ‘depressive disorder’ OR ‘major depressive disorder’ OR ‘clinical depression’ OR ‘mood disorder’)

#### AND

(2018:py OR 2019:py OR 2020:py OR 2021:py OR 2022:py OR 2023:py OR 2024:py) AND [embase]/lim NOT ([embase]/lim AND [medline]/lim) AND (’article’/it OR ‘preprint’/it)

#### Scopus

((“Large Language Model” [all AND Fields] OR “LLM” [all AND Fields] OR “Generative Pre-trained Transformer” [all AND Fields] OR “GPT” [all AND Fields] OR “GPT-2” [all AND Fields] OR “GPT-3” [all AND Fields] OR “GPT-3.5” [all AND Fields] OR “GPT-4” [all AND Fields] OR “ChatGPT” [all AND Fields] OR “Transformer models” [all AND Fields] OR “BERT” [all AND Fields] OR “BARD” [all AND Fields] OR “Gemini” [all AND Fields]) AND (“Depression” [all AND Fields] OR “Depressive disorder” [all AND Fields] OR “Major depressive disorder” [all AND Fields] OR “Clinical depression” [all AND Fields] OR “Mood disorder” [all AND Fields])) AND (PUBYEAR > 2017 AND PUBYEAR < 2025) AND (LIMIT-TO (DOCTYPE, “ar”))

#### Web of science

(TS=(“Large Language Model” OR “LLM” OR “Generative Pre-trained Transformer” OR “GPT” OR “GPT-2” OR “GPT-3” OR “GPT-3.5” OR “GPT-4” OR “ChatGPT” OR “Transformer models” OR “BERT” OR “BARD” OR “Gemini”) AND TS=(“Depression” OR “Depressive disorder” OR “Major depressive disorder” OR “Clinical depression” OR “Mood disorder”)) AND PY=2018-2024

#### Risk of bias

**Table S1:**
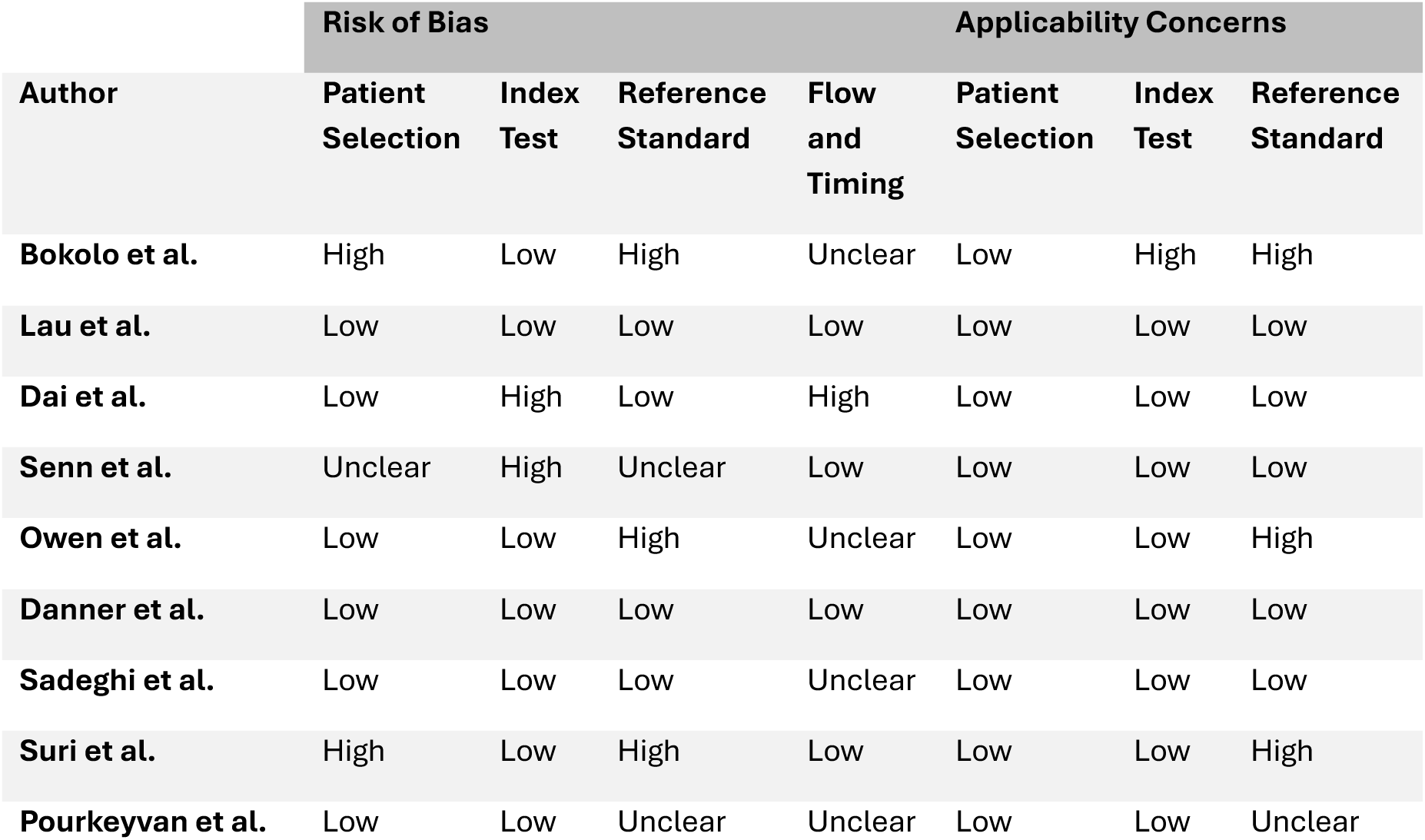
The results of the risk of bias assessment according to the Quality Assessment of Diagnostic Accuracy Studies 2 (QADAS-2) tool.

**Table S2:**
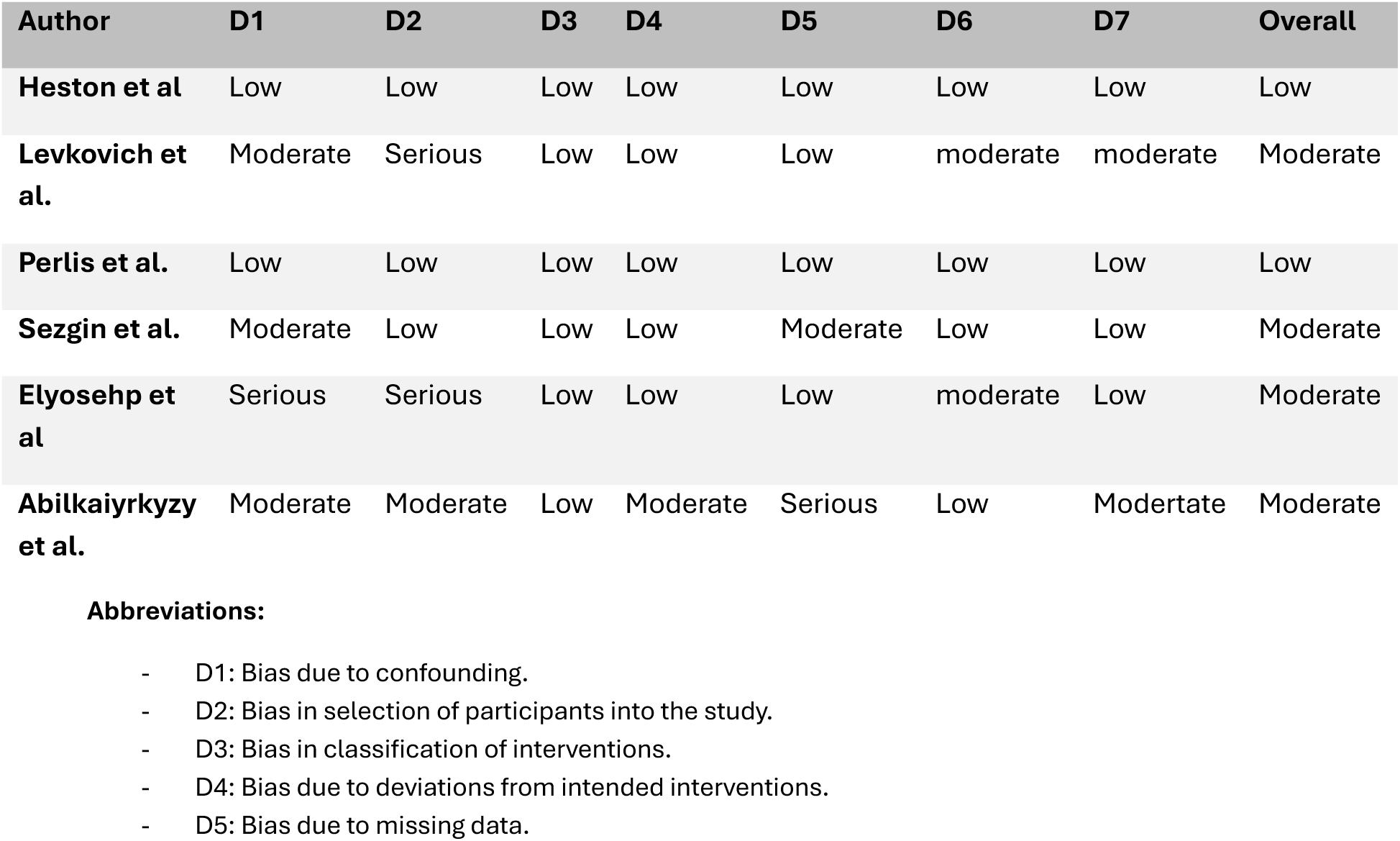
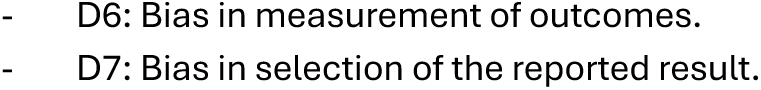
The results of the risk of bias assessment according to the Risk Of Bias In Non-randomized Studies - of Interventions (ROBINS-I) tool.

| Author | D1 | D2 | D3 | D4 | D5 | D6 | D7 | Overall |
| --- | --- | --- | --- | --- | --- | --- | --- | --- |
| Heston et al | Low | Low | Low | Low | Low | Low | Low | Low |
| Levkovich et al. | Moderate | Serious | Low | Low | Low | moderate | moderate | Moderate |
| Perlis et al. | Low | Low | Low | Low | Low | Low | Low | Low |
| Sezgin et al. | Moderate | Low | Low | Low | Moderate | Low | Low | Moderate |
| Elyosehp et al | Serious | Serious | Low | Low | Low | moderate | Low | Moderate |
| Abilkaiyrkyzy et al. | Moderate | Moderate | Low | Moderate | Serious | Low | Modertate | Moderate |

**Table S3:** The results of the risk of bias assessment according to the Prediction model Risk Of Bias ASsessment Tool (PROBAST) tool.

| Author | Risk of bias |  |  |  | Applicability |  |  |
| --- | --- | --- | --- | --- | --- | --- | --- |
|  | Participants | Predictors | Outcome | Analysis | Participants | Predictors | Outcome |
| Wan et al. | high | low | low | high | high | low | low |
| Wang et al. | Low | Unclear | High | Unclear | Low | Low | Low |
| Hond et al. | Low | low | low | Low | Low | Low | Low |
| Danner et al. | Low | High | High | Low | High | Low | Low |
| Farruque et al. | Low | low | low | Low | Low | Low | Low |
| Lu et al. | Low | Unclear | low | Low | Low | Unclear | Low |
| Lam et al. | Low | low | low | Low | Low | Low | Low |
| LLias et al. | Low | low | low | Low | Low | Low | Low |
| Toto et al. | Low | low | low | Low | Low | Low | Low |
| Kabir et al. | Low | Low | Unclear | Low | Low | Low | Low |
| Farruque et al. | High | Low | Low | Low | High | Low | Low |
| Janatdoust et al. | High | Low | Low | Low | High | Low | Low |
| Adarsh S et al. | High | Low | Low | Low | High | Low | Low |
| Sivamanikandan S. et al. | High | Low | Low | Low | High | Low | Low |
| Esackimuthu et al. | High | Low | Low | Unclear | High | Low | Low |
| Singh et al. | High | Low | Low | Low | High | Low | Low |
| Poświata et al. | High | Low | Low | Low | High | Low | Low |
| Hegde et al. | High | Low | Low | Low | High | Low | Low |

