## Supplementary material for "Exploring the Efficacy and Potential of Large Language Models for Depression: A Systematic Review": Tables included in the manuscript

**Table 1: Summary of included studies**

| **Author** (Ref) | **Year** | **Sample Size & Data Type** | **Task Type** | **Specific Task** | **Summary of Key Results** |
|---|---|---|---|---|---|
| **Bokolo et al. (31)** | 2023 | 632,000 tweets | Detection | Depression detection from Twitter | RoBERTa achieved high accuracy; transformer models outperformed ML models. |
| **Lau et al. (26)** | 2023 | 189 clinical interview transcripts | Classification | Depression severity assessment | Prefix-tuned LLMs outperformed traditional models with lower error rates. |
| **Dai et al. (32)** | 2021 | 500 EHRs | Classification | Psychiatric patient screening | Transfer learning models like DistilBERT and ROBERTa improved performance. |
| **Heston et al. (51)** | 2023 | 25 conversational agents | Detection and Management | Depression and suicidality detection | Agents recommended intervention late; delayed suicide hotline reference. |
| **Senn et al. (40)** | 2022 | 189 clinical interviews | Classification | Depression classification from interviews | BERT ensembles showed robustness and higher F1 scores. |
| **Levkovich et al. (7)** | 2023 | 8 vignettes | Classification and Management | Treatment strategy assessment | ChatGPT aligned with treatment guidelines, differing from primary care physicians. |
| **Perlis et al. (29)** | 2024 | 50 clinical vignettes | Management | Bipolar depression management | Augmented LLMs matched expert treatment choices better than non-augmented models. |
| **Sezgin et al. (28)** | 2023 | 14 PPD questions | Management | Postpartum depression information quality assessment | ChatGPT provided more clinically accurate responses than other models. |

| | | | | | |
|---|---|---|---|---|---|
| **Wan et al. (44)** | 2022 | 12,006 admission notes | Classification | Family history identification in mood disorders | BERT–CNN model achieved high accuracy in family history extraction. |
| **Owen et al. (39)** | 2023 | Reddit datasets | Detection | Depression signal detection in Reddit posts | BERT and MentalBERT showed significant early detection ability. |
| **Wang et al. (35)** | 2020 | 13,993 microblogs | Classification | Depression risk prediction from Weibo posts | BERT achieved the highest micro-averaged F1 score. |
| **Elyosehp et al. (3)** | 2024 | Case vignettes | Classification and Management | Depression prognosis assessment | LLMs closely aligned with human professionals in prognosis. |
| **Hond et al. (50)** | 2024 | 16,159 cancer patients' EHR data | Detection | Early depression risk detection in cancer patients | Structured EHR data used alone yielded the best prediction results. |
| **Danner et al. (25)** | 2023 | DAIC-WOZ datasets | Detection | Depression detection from interviews | Models achieved superior performance, significantly outperforming others. |
| **Farruque et al. (36)** | 2024 | 6077 tweets and 1500 annotated tweets | Detection and Classification | Depression symptoms modelling from Twitter | Iterative LLM training improved accuracy in depression detection. |
| **Lu et al. (34)** | 2023 | DAIC dataset | Detection | Depression detection via conversation turn classification | Deep learning framework achieved a high F1-score. |
| **Lam et al. (30)** | 2019 | 189 DAIC-WOZ participants | Detection | Multi-modal depression detection | Multi-modal model outperformed single-modality in F1 score. |

| | | | | | |
|---|---|---|---|---|---|
| **Llias et al. (27)** | 2023 | Public datasets | Detection | Stress and depression identification in social media | Enhanced performance with multimodal adaptation gates. |
| **Toto et al. (24)** | 2021 | 189 clinical interviews | Detection | Depression screening using audio and text | AudiBERT significantly improved classification with high F1 scores. |
| **Sadeghi et al. (41)** | 2023 | 275 interview transcripts | Detection | Depression severity prediction from text | Fine-tuned GPT-3.5-Turbo showed robust accuracy. |
| **Kabir et al. (37)** | 2023 | 40,191 tweets | Detection and Classification | Depression severity detection from tweets | Attention-based models classified tweets with high confidence scores. |
| **Suri et al. (43)** | 2022 | 5997 tweets | Detection | Depressive tendencies detection using multimodal data | Cross-modal attention-based BERT model showed high F1 scores. |
| **Abilkaiyrkyzy et al. (38)** | 2024 | 219 E-DAIC participants | Detection and Management | Mental illness detection using a chatbot | Chatbot achieved 69% accuracy and high usability. |
| **Pourkeyvan et al. (42)** | 2024 | 11,890,632 tweets and 553 bio-descriptions | Detection | Mental health disorder prediction from Twitter | Hugging Face BERT models significantly improved prediction accuracy. |
| **Tey et al. (49)** | 2023 | Over 3.5 million tweets | Detection and Classification | Pre- and post-depressive detection from tweets | BERT with emoji decoding accurately classified depressive categories. |
| **Farruque et al. (33)** | 2022 | 13,387 Reddit samples | Detection and Classification | Depression level detection modelling | MBERT improved classification, especially for longer posts. |
| **Janatdoust et al. (46)** | 2022 | 16,632 social media comments | Detection and Classification | Depression signs detection from social media text | BERT-based ensembles demonstrated superior classification accuracy. |

| | | | | | |
|---|---|---|---|---|---|
| **Adarsh S et al. (54)** | 2022 | Social media texts | Detection and Classification | Depression sign detection using BERT | Fine-tuned BERT model showed specificity in detection improvement. |
| **Sivamanikandan S. et al. (52)** | 2022 | Social media posts | Detection and Classification | Depression level classification | RoBERTa achieved best performance among the tested transformer models. |
| **Esackimuthu et al. (53)** | 2022 | ALBERT base v1 data | Detection and Classification | Depression detection from social media text | ALBERT model showed potential despite challenges with data quality. |
| **Singh et al. (45)** | 2022 | Ensemble of models | Detection and Classification | Depression level classification using BERT, RoBERTa, XLNet | Ensemble ranked 3rd, indicating effective detection and classification. |
| **Poświata et al. (48)** | 2022 | RoBERTa models' data | Detection and Classification | Depression sign detection using RoBERTa | Ensemble achieved high performance metrics in competitive setting. |
| **Hegde et al. (47)** | 2022 | Social media text data | Detection and Classification | Depression detection using supervised learning | BERT-based TL model performed better than ML classifiers. |

**Abbreviations**: Ref: Reference | EHR: Electronic Health Record | LLM: Large Language Model | NLP: Natural Language Processing | PHQ-8: Patient Health Questionnaire-8 | DAIC-WOZ: Distress Analysis Interview Corpus - Wizard of Oz | CNN: Convolutional Neural Network | BERT: Bidirectional Encoder Representations from Transformers | AUROC: Area Under the Receiver Operating Characteristic Curve | ZSL: Zero-Shot Learning | TL: Transfer Learning | MLP: Multi-Layer Perceptron | ACOG: American College of Obstetricians and Gynecologists | DSM-5: Diagnostic and Statistical Manual of Mental Disorders, Fifth Edition | SVM: Support Vector Machine | BiLSTM: Bidirectional Long Short-Term Memory | ROC AUC: Receiver Operating Characteristic Area Under the Curve.

**Table 2: Summary of conclusions and limitations of included studies**

| Author | Model | Task Type | Conclusion Summary | Limitations Summary |
|---|---|---|---|---|
| **Bokolo et al. (31)** | RoBERTa, DeBERTa | Detection | Transformer models like RoBERTa excel in depression detection from Twitter data, outperforming traditional ML approaches. | Dataset not initially intended for depression detection, lacks demographic and linguistic diversity. |
| **Lau et al. (26)** | Prefix-tuned LLM | Classification | LLMs with prefix-tuning significantly enhance depression severity assessment, surpassing traditional methods. | Challenges include potential overfitting, limited training data, and lack of generalizability. |
| **Dai et al. (32)** | BERT, DistilBERT, ALBERT, ROBERTa | Classification | BERT models, especially with feature dependency, effectively classify psychiatric conditions from EHRs. | Highly imbalanced dataset and overlapping psychiatric symptoms complicate accurate classification. |
| **Heston et al. (51)** | GPT-3.5 | Detection and Management | Conversational agents show delayed response in escalating mental health risks, needing more rigorous testing. | Limited by the use of publicly available agents and structured prompts, lacking real-world interaction complexity. |
| **Senn et al. (40)** | BERT, RoBERTa, DistilBERT | Classification | Ensembles of BERT models enhance depression detection robustness in clinical interviews. | Small dataset size and reliance solely on transcript data limit effectiveness. |
| **Levkovich et al. (7)** | GPT-3.5 and GPT-4 | Classification and Management | ChatGPT models align with treatment guidelines better than primary care physicians in hypothetical scenarios. | Study based on vignettes may not accurately reflect real patient interactions. |
| **Perlis et al. (29)** | GPT-4 | Management | Augmented GPT-4 aids in clinical decision support for bipolar disorder, outperforming non-augmented models. | Limited by the narrow scope of clinical vignettes used, questioning the generalization of findings. |
| **Sezgin et al. (28)** | GPT-4, LaMDA | Management | GPT-4 provides more accurate responses to postpartum depression queries than other models and traditional searches. | Reliance on a limited set of standardized questions and the non-medical focus of LLM design limit applicability. |

| | | | | |
|---|---|---|---|---|
| **Wan et al. (44)** | BERT–CNN | Classification | High accuracy in identifying family psychiatric history from EHRs, suggesting utility in understanding mood disorders. | Study's applicability is limited to a single hospital's dataset, possibly not generalizable. |
| **Owen et al. (39)** | BERT, MentalBERT | Detection | Effective identification of depressive signals in online forums, with potential for early intervention. | Challenges in determining exact timing of posts and interpreting informal internet communication. |
| **Wang et al. (35)** | BERT, RoBERTa, XLNET | Classification | Deep learning methods, enhanced by domain-specific pretraining, effectively detect depression risk levels from microblogs. | Data imbalance and semantic ambiguities in microblogs complicate accurate depression risk classification. |
| **Elyosehp et al. (3)** | GPT-3.5, GPT-4, Claude, Bard | Classification and Management | LLMs match or surpass professional judgments in prognosis accuracy, showing potential for clinical integration. | Vignette-based methodology limits real-world applicability; further validation with actual patient data is needed. |
| **Hond et al. (50)** | BERT | Detection | Machine learning models predict depression risk in cancer patients using EHRs, with structured data models performing best. | Bias in model calibration and reliance on structured data might miss unrecorded symptoms. |
| **Danner et al. (25)** | BERT-based models, GPT-3.5, GPT-4 | Detection | Advanced transformer networks significantly enhance depression detection from clinical interview data. | Ethical, legal, and privacy concerns need addressing; further validation required. |
| **Farruque et al. (36)** | BERT, Mental-BERT | Detection and Classification | Semi-supervised learning models, iteratively refined with Twitter data, improve depression symptom detection accuracy. | Limited dataset size and absence of continuous human annotation during model training may affect reliability. |
| **Lu et al. (34)** | BERT, transformer encoder | Detection | Novel deep learning framework enhances depression detection from psychiatric interview data, improving interpretability. | Study limited to transcribed data, which may not capture all nuances of psychiatric assessments. |
| **Lam et al. (30)** | Transformer, 1D CNN | Detection | Multi-modal models combining text and audio data effectively detect depression, enhanced by data augmentation. | Limited generalizability due to dataset specificity and potential biases in class distribution. |

| | | | | |
|---|---|---|---|---|
| **Llias et al. (27)** | BERT, MentalBERT | Detection | Extra-linguistic features improve calibration and performance of models in detecting stress and depression from texts. | Constraints in GPU resources, lack of explainability, and reliance on single model runs limit robustness. |
| **Toto et al. (24)** | AudiBERT | Detection | AudiBERT outperforms traditional and hybrid models in depression screening, utilizing multimodal data. | Emotional expression variability and privacy concerns during data collection impact model training and performance. |
| **Sadeghi et al. (41)** | GPT-3.5-Turbo, DepRoBERTa | Detection | Language models effectively predict depression severity from textual data, enhancing diagnostic procedures. | Limited dataset size impacts generalizability of findings. |
| **Kabir et al. (37)** | BERT, DistilBERT | Detection and Classification | Models effectively classify social media texts into depression severity categories, with high confidence and accuracy. | Annotation biases and lack of contextual understanding in social media texts pose significant challenges. |
| **Suri et al. (43)** | BERT | Detection | Multimodal BERT frameworks significantly enhance detection of depressive tendencies from complex social media data. | Data biases and model applicability to less active or non-openly expressive users are noted concerns. |
| **Abilkaiyrkyzy et al. (38)** | BERT | Detection and Management | Chatbot effectively detects and classifies mental health issues, highly usable for reducing barriers to mental health care. | Chatbot's limited emotional detection capabilities and dataset specificity restrict broader applicability. |
| **Pourkeyvan et al. (42)** | BERT models from Hugging Face | Detection | Superior detection of depression symptoms from social media, demonstrating the efficacy of advanced NLP models. | Analysis limited to English-language tweets, not representing non-English speaking populations. |
| **Tey et al. (49)** | BERT, supplemented with emoji decoding | Detection and Classification | Augmented BERT model classifies Twitter users into depressive categories, enhancing early depression detection. | Reliance on self-reported diagnosis and English-only analysis limits generalizability. |

| | | | | |
|---|---|---|---|---|
| **Farruque et al. (33)** | Mental BERT (MBERT) | Detection and Classification | MBERT enhanced with text excerpts significantly improves depression level classification from social media posts. | High computational demands for excerpt extraction limit practical application in time-sensitive environments. |
| **Janatdoust et al. (46)** | Ensemble of BERT, ALBERT, DistilBERT, RoBERTa | Detection and Classification | Ensemble models effectively classify depression signs from social media, utilizing multiple language models for improved accuracy. | Training based on predefined criteria may not accurately reflect complex depressive symptoms. |
| **Adarsh S et al. (54)** | BERT-small | Detection and Classification | Enhanced BERT model accurately classifies depression severity from social media texts, understanding nuances better than others. | Natural language variability and contextual depth of posts, alongside imbalanced data classes, pose challenges. |
| **Sivamanikandan S. et al. (52)** | DistilBERT, RoBERTa, ALBERT | Detection and Classification | Transformer models classify depression levels effectively, with RoBERTa achieving the best performance. | Dataset imbalance and lack of clinical validation limit the generalization of findings. |
| **Esackimuthu et al. (53)** | ALBERT base v1 | Detection and Classification | ALBERT shows potential in detecting depression signs from social media texts but faces challenges due to complex human emotions. | Limited by the quality of social media data and subtle nature of human emotions. |
| **Singh et al. (45)** | Ensemble of BERT, RoBERTa, XLNet | Detection and Classification | Ensemble model accurately classifies depression levels from social media text, ranking highly in competitive settings. | Reliance on social media text and absence of real-world clinical validation pose significant challenges. |
| **Poświata et al. (48)** | RoBERTa, DepRoBERTa | Detection and Classification | RoBERTa and DepRoBERTa ensemble excels in classifying depression signs, securing top performance in a competitive environment. | Limited by dataset specifics and the competitive model training environment. |

| | | | | |
|---|---|---|---|---|
| **Hegde et al. (47)** | Ensemble of ML classifiers, BERT | Detection and Classification | BERT-based Transfer Learning model outperforms traditional ML classifiers in detecting depression from social media texts. | Challenges include handling the natural language variability and contextual depth of social media posts. |

**Abbreviations**: BERT: Bidirectional Encoder Representations from Transformers | CNN: Convolutional Neural Network | GPT: Generative Pre-trained Transformer | LLM: Large Language Model | MLP: Multi-Layer Perceptron | NLP: Natural Language Processing | PHQ-8: Patient Health Questionnaire-8 | PHQ-9: Patient Health Questionnaire-9 | TL: Transfer Learning | ZSL: Zero-Shot Learning.
